# Comparative Evaluation of Pretrained Large Language Models for Suicide Risk Prediction from Clinical Notes in U.S. Veterans

**DOI:** 10.64898/2026.06.16.26355804

**Authors:** Joshua Levy, Maxwell Levis, Monica Dimambro, Luke Rozema, Siamack Ayandeh, Alos Diallo, Yefan Zhou, Siting Li, Weiyi Wu, Brian Shiner, Jiang Gui

## Abstract

**Background:** Suicide remains a significant and potentially preventable cause of death among United States veterans. Predictive models based on structured electronic health record (EHR) data, including the U.S. Department of Veterans Affairs’ Recovery Engagement and Coordination for Health–Veterans Enhanced Treatment (REACH-VET) program, aim to identify individuals at elevated risk for enhanced monitoring and follow-up. Increasing evidence suggests that unstructured clinical narratives contain additional psychosocial information that may enhance risk prediction when analyzed using natural language processing (NLP). However, optimal approaches for representing clinical text remain uncertain. Recent advances in large language models (LLMs) enable contextual text representations that capture complex semantic relationships beyond traditional lexical methods.

**Methods:** We compared the predictive performance of pretrained LLMs with classical bag-of-words (BoW) representations for suicide risk prediction using clinical notes from 27,241 veterans receiving care in the Veterans Health Administration. Patients were stratified by REACH-VET risk tier (low, moderate, high), and models were evaluated across prediction windows defined by note look-back periods (<30, <90, and <270 days).

**Results:** LLM-based representations outperformed BoW approaches in seven of nine risk tier–time window combinations, achieving a maximum AUROC of 0.644 when solely considering text. Incorporating structured clinical variables further improved performance (AUROC=0.748). Model interpretation identified suicide-related language, especially in notes documented within 30 days of the outcome among patients classified as high risk.

**Conclusions:** Pretrained LLMs can extract clinically meaningful information from narrative documentation, providing a foundation for future work adapting to additional clinical contexts and nuanced temporal associations to improve suicide risk prediction.

## Introduction

Suicide remains a critical public health concern among U.S. veterans, whose rates far exceed those observed in civilian populations ^1^ . In response, the U.S. Department of Veterans Affairs (VA) has made suicide prevention a top priority, investing in tools that enable early detection and intervention ^2^. A cornerstone of this effort is the Recovery Engagement and Coordination for Health – Veterans Enhanced Treatment (REACH-VET) program, which applies predictive algorithms to structured electronic health record (EHR) data to identify individuals at elevated suicide risk ^3^. By flagging high-risk patients, the VA can direct prevention resources toward those most in need.

Recent research has demonstrated that supplementing structured EHR data with unstructured sources, such as clinical notes, can enhance the accuracy of suicide risk models ^4^. In earlier work, we leveraged natural language processing (NLP) techniques to analyze these narratives, identifying linguistic variables that enriched traditional demographic and structured predictors across different risk tiers ^5^. These findings underscore the potential of text-derived features to reveal nuanced clinical signals that structured data alone may overlook.

Although using dictionary derived semantic indexes may help track established psychosocial risk constructs ^6^, relying exclusively on such databases has notable limitations. Predictive lexical patterns relevant to suicide may exist outside predefined semantic categories. Prior studies have often addressed this gap using bag-of-words approaches, which model clinical notes by counting word frequencies ^7^. However, bag-of-words representations treat each word as an isolated feature, resulting in sparse, high-dimensional data and ignoring important contextual information. Word embedding methods address these challenges by mapping words into dense, low-dimensional vector spaces (i.e., embeddings) that encode relationships between terms based on their co-occurrence patterns ^8^. This approach allows entire clinical notes to be represented as ordered sequences of vectors, capturing contextual meaning while reducing sparsity and computational burden.

Deep learning approaches use multi-layered artificial neural network algorithms to improve on text embeddings to learn complex patterns in clinical language. Rather than treating words independently, these models learn how words relate to one another within a sentence and across an entire document, allowing them to form increasingly abstract and informative representations of text. Some approaches focus on word order to capture how meaning unfolds over time ^9^, while others use attention mechanisms to highlight the most relevant parts of a note and integrate information from across the full context ^10^. Benchmark studies consistently show that deep learning methods outperform traditional lexical approaches for various predictive tasks by identifying recurrent syntactic and semantic patterns across documents ^11,12^. This ability to distill rich contextual information into compact, predictive representations offers a powerful foundation for improving suicide risk prediction models.

The emergence of large language models (LLMs) built on the transformer architecture represents a major advancement in natural language processing ^10^. Examples of LLMs include BERT (Bidirectional Encoder Representations from Transformers), Generative Pretraining (GPT), BigBird, and Longformer ^13–16^. These models use self-attention mechanisms to consider relationships among all words in a sentence or document simultaneously, enabling the contextual meaning of each word to be interpreted in relation to all others. Unlike traditional NLP methods that rely on predefined features, transformer architectures learn directly from raw text, preserving both local and global dependencies. By producing dense, context-rich embeddings of entire clinical narratives, these models offer state-of-the-art tools for analyzing sequential and longitudinal text data, making them particularly promising for suicide risk prediction tasks that require subtle interpretation of language patterns. Although recent generative AI approaches can evaluate or summarize clinical reports using rubric-based prompts, the present study focuses on encoder-based models that convert clinical notes into numerical representations for risk prediction. This approach allows clinical narratives to be systematically integrated with structured patient data for downstream predictive modeling.

In this study, we compare the predictive performance of LLM encoders with that of traditional count-based approaches for suicide risk prediction in a large veteran population. As a proof of concept, we employ large language models without corpus-specific fine-tuning to establish a performance baseline. This research lays the groundwork for future efforts aimed at refining these models to capture time-sensitive linguistic signatures associated with suicide risk.

## Methods

**Figure 1.**
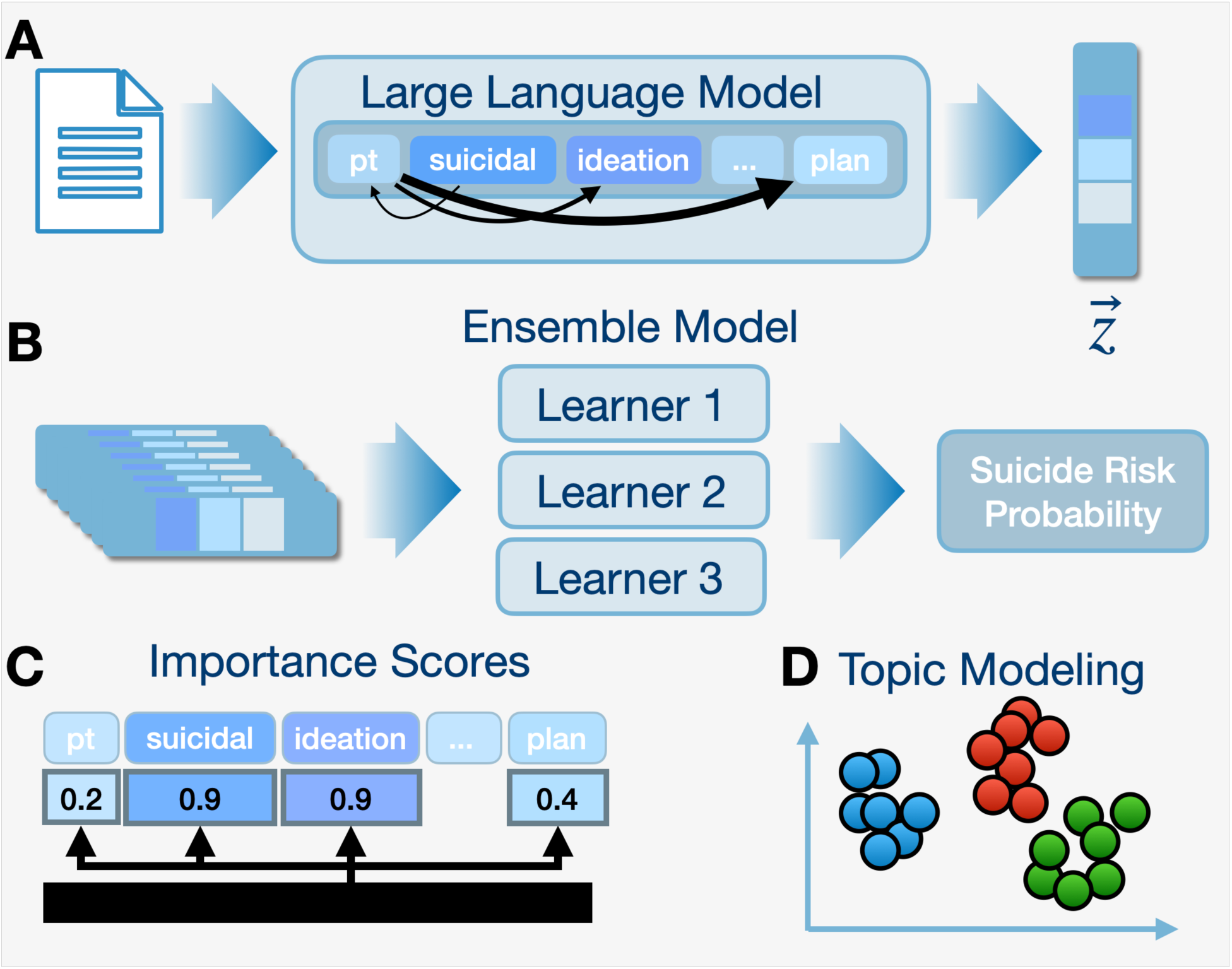
Overview of the suicide risk prediction and interpretation framework. **A)** Clinical notes are encoded using a pretrained large language model to generate dense contextual embeddings (***z***). **B)** These embeddings are passed into a gradient-boosted ensemble classifier to estimate patient-level suicide risk probabilities. **C)** Model interpretability is assessed at multiple levels, including word-level contribution visualizations and corpus-level analyses using low-dimensional embedding projections (e.g., UMAP) to identify **D)** semantically coherent clusters of clinical language associated with suicide risk.

### Study Cohort Description and Extraction of Structured Patient Characteristics

Building on earlier methodological frameworks, we assembled a retrospective matched case-control cohort using data from the U.S. Department of Veterans Affairs (VA) Corporate Data Warehouse (CDW), a national repository of electronic health records, supplemented with mortality information from the VA-Department of Defense Mortality Data Repository. Veterans who had received VA healthcare services during 2017–2018 and subsequently died by suicide were identified, resulting in 4,584 case patients. Each case was matched to five control participants who were alive on the index date (the case’s date of death), producing 22,657 matched controls (**Table 1**). Matching criteria included VA facility, calendar time, and suicide risk percentile, based on REACH-VET, the VA’s leading suicide risk prediction algorithm. site of care or baseline clinical risk and to facilitate comparison with the REACH-VET risk model. Matching was done to increase sample size, statistical power ^17^, and reduce confounding from differences in site of care, baseline clinical risk, and facilitate comparison with the REACH-VET risk model.

**Table 1.** Patient Characteristics for parent 2017-2018 cohort with clinical notes extracted from one year time to death (used to derive 30, 90 and 270 day subcohorts); omitted variables relating to eligibility, disability, physical/mental health burden, select mental health risks, inpatient/ER, select prescriptions

|  | Case<br>(N=4584) | Control<br>(N=22657) | SMD |
| --- | --- | --- | --- |
| **Demographics** |  |  |  |
| Female | 163 (3.6%) | 1565 (6.9%) | 0.151 |
| Married | 1931 (42.1%) | 10268 (45.3%) | 0.064 |
| Homeless Prior 2 Years | 60 (1.3%) | 318 (1.4%) | 0.008 |
| Veteran | 4542 (99.1%) | 22495 (99.3%) | 0.022 |
| Rural | 60 (1.3%) | 318 (1.4%) | 0.006 |
| Age: Mean (SD) | 61.0 (18.5) | 60.6 (16.5) | 0.024 |
| **Race** |  |  |  |
| Native American / Asian / Pacific Islander | 101 (2.3%) | 505 (2.2%) | 0.259 |
| Black | 240 (5.2%) | 2399 (10.6%) |  |
| Hispanic | 184 (4.0%) | 1355 (6.0%) |  |
| White | 3774 (82.3%) | 17692 (78.1%) |  |
| **Risk Tier** |  |  |  |
| High | 451 (10.2%) | 2205 (10.2%) | 0.948 |
| Medium | 2091 (47.4%) | 10306 (47.7%) |  |
| Low | 2042 (42.4%) | 10146 (42.1%) |  |
| **Deployment** |  |  |  |
| Vietnam | 1671 (36.5%) | 9167 (40.5%) | 0.084 |
| Afghanistan or Iraq | 1541 (33.6%) | 7931 (35.0%) | 0.037 |
| **Mental Health Risk Flag** |  |  |  |
| Anxiety | 1835 (40.0%) | 9187 (40.5%) | 0.011 |
| Bipolar | 725 (15.8%) | 2841 (12.5%) | 0.094 |
| Depression | 2649 (57.8%) | 12744 (56.2%) | 0.031 |
| PTSD | 1451 (31.7%) | 7161 (31.6%) | 0.001 |
| Sleeping | 1865 (40.7%) | 10023 (44.2%) | 0.073 |
| Substance | 1779 (38.8%) | 7374 (32.5%) | 0.131 |
| Trauma | 1985 (43.3%) | 9986 (44.1%) | 0.016 |
| **Prescriptions Prior 12 Months** |  |  |  |
| Opioid Rx | 1073 (23.4%) | 5202 (23.0%) | 0.011 |
| Mood Stabilizer Rx | 1263 (27.6%) | 5835 (25.8%) | 0.041 |
| Antipsychotic Rx | 748 (16.3%) | 2838 (12.5%) | 0.108 |
| Antidepressant Rx | 2000 (43.6%) | 9887 (43.6%) | 0.000 |

Structured variables were drawn from the VA CDW to characterize each patient’s demographic, clinical, and healthcare utilization profile. Extracted variables encompassed demographic attributes such as age, sex, race or ethnicity, marital status, and military service branch, as well as indicators of social vulnerability including prior experiences of homelessness. Measures of healthcare utilization captured the frequency and types of encounters across settings, including primary care, emergency, specialty, and mental health services. Medication exposure data were compiled from pharmacy dispensing records and included prescriptions for psychotropic and pain-related medications, such as opioids, antipsychotics, and antidepressants. Diagnostic information was derived from ICD-coded problem lists and encounter diagnoses, covering a broad spectrum of mental health and substance use disorders, including posttraumatic stress disorder, depression, anxiety, and bipolar disorder. To address missing data (only 2.4% of patients were missing at least five of the 61 variables), we used multivariate imputation by chained equations (MICE) ^18^. Following data cleaning and harmonization, 61 structured variables were available for analysis. These structured clinical variables, summarized in **Table 1**, complemented the unstructured clinical text used in downstream modeling.

### Risk Stratification and Note Selection

To account for heterogeneity in baseline suicide risk, patients were grouped into three strata corresponding to previously defined REACH-VET risk tiers: low (REACH-VET risk scores from 100–25), moderate (24–2), and high (1) ^4,19^. For each tier, unstructured clinical notes were extracted for three look-back intervals (‘time windows’): 30, 90, and 270 days preceding the index date (or the corresponding date for controls) ^4,20^. Patients were included in each interval only if they had at least one note within that time window; individuals lacking notes in shorter intervals were excluded from those subsets.

To minimize bias associated with documentation immediately preceding death, we applied a more stringent exclusion criterion than in earlier work– all notes dated within six days of the index date were removed ^20^. This conservative window helped ensure that models were trained on clinical text reflective of ongoing care rather than acute crisis documentation or potentially endogenous information reporting suicide events.

Patients with unusually high documentation volume, defined as more than six times the average number of notes within a given interval, were excluded to prevent model bias toward high utilizers. After applying all inclusion and exclusion criteria, the final dataset consisted of three strata: (1) low-risk tier patients (378,697 notes from 1,934 cases and 9,702 controls), (2) moderate-risk tier patients (859,075 notes from 2,055 cases and 10,105 controls), and (3) high-risk tier patients (566,710 notes from 449 cases and 2,198 controls).

Clinical text underwent a sequence of conventional preprocessing operations prior to feature extraction. All text was converted to lowercase, punctuation was stripped, and frequently occurring function words (e.g., “the”, “and”, “was”) were removed to reduce noise. A note deidentification protocol was developed, validated and applied to the study cohort.

### Clinical Note Bag-of-Words Representations

For generating data that could be utilized for the Bag-of-Words (BoW) approaches, cleaned texts were segmented into individual tokens and expanded to include both unigrams and bigrams. Feature matrices were generated using Scikit-learn ^21^, applying both term frequency–inverse document frequency (TF–IDF) transformations and raw token counts. To manage vocabulary size and limit sparsity, terms were filtered based on document frequency: only those appearing in at least 1% but no more than 90% of documents within each risk tier and time window were retained.

### Clinical Text Feature Extraction Using Large Language Model Embeddings

We utilized several pretrained LLMs to convert tokenized clinical text into numerical vector embeddings that capture the semantic and contextual information contained in each note (**Figure 1A**). These models were used in an off-the-shelf (zero-shot; without additional fine-tuning) configuration because their pretraining on extensive biomedical and clinical corpora was expected to sufficiently represent the linguistic characteristics of VA clinical notes ^22^. This approach ensured a fair comparison with BoW methods, serving as an initial baseline for future fine-tuned model development. The decision to use pretrained models and their respective model architectures without further optimization was also guided by practical considerations, including privacy-preserving GPU resource limitations that constrained large-scale fine-tuning of large parameter models at the time of analysis.

All models were implemented through the Hugging Face Transformers library ^23^. The following pretrained architectures were employed:

1. **BioClinicalBERT** – A domain-specific variant of BERT pretrained on biomedical literature (PubMed abstracts and full-text articles) and further refined using over two million clinical notes from the MIMIC-III intensive care database at Beth Israel Deaconess Medical Center ^14,24^. BioClinicalBERT uses a bidirectional encoder structure with multi-head self-attention layers, enabling each word’s meaning to be informed by its surrounding context. The model can process a maximum of 512 tokens per input sequence. Longer notes were segmented into overlapping 512-token chunks, and resulting embeddings were averaged across segments to generate a single note-level representation.
2. **Clinical Longformer** – A transformer architecture optimized for long-sequence input, extending the maximum input length from 512 to 4,096 tokens through the use of sliding window and sparse attention mechanisms. This allows full-document processing while maintaining computational efficiency. Clinical Longformer was also pretrained on the MIMIC-III dataset ^15,16^.
3. **Clinical BigBird** – Similar in purpose to Longformer, BigBird additionally uses random attention patterns to improve the capture of broader contextual relationships across lengthy clinical narratives ^16^.

These models are widely used in contemporary biomedical NLP research for benchmarking and comparative evaluation ^22^. Because these pretrained architectures lack a task-specific classification token (CLS), we applied mean pooling across the final hidden layer to derive a fixed-size note embedding. Each clinical note was thus represented by a 768-dimensional vector (output dimensionality of the penultimate neural network layer) summarizing its overall contextual and semantic content.

### Prediction of Suicide Risk and Integration of Patient Characteristics

Gradient-boosted tree models (XGBoost) ^25^ were developed to classify suicide decedents within each REACH-VET risk tier and temporal window (<30, <90, and <270 days) (**Figure 1B**). Two primary feature sets were evaluated: one derived from BoW representations and another from zero-shot LLM embeddings. Separate models were trained for each feature configuration to enable direct comparison of text representation approaches.

Model training followed a standard machine-learning workflow, dividing notes into training, validation, and test sets (64%-16%-20% splits). Hyperparameters were optimized using a randomized grid search, with early stopping applied using the validation set to prevent overfitting. All training and interpretation were conducted at the note level, producing a predicted probability for each clinical note. For patient-level evaluation, note-level predictions were aggregated by averaging probabilities to yield a single score representing overall suicide risk. Model selection was guided by the validation-set area under the receiver operating characteristic curve (AUROC). Final model performance was assessed on a held-out test set, and 95% confidence intervals for AUROC were estimated using 1,000 non-parametric bootstrap resamples with replacement.

To examine the contribution of structured patient characteristics, four configurations were tested: Bag of Words (BoW)-only (no patient characteristics), LLM-only, BoW plus structured variables (BoW+Characteristics), and LLM plus structured variables (LLM+Characteristics). Structured data, including demographics, healthcare utilization, and diagnostic variables, were concatenated to note-level embeddings for the combined models prior to the aggregation strategy. Because the number of text features greatly exceeded the number of structured predictors, a sampling adjustment was applied during training to upweight structured features using a methodology established in a prior work ^6^, ensuring more balanced model learning across data modalities.

### Model Interpretation

Interpretation of the LLM outputs was performed at two levels: *note-level* and *corpus-level*. These complementary analyses were designed to identify salient linguistic features and contextual themes associated with suicide risk. Note-level interpretation was conducted on a random subset of notes from the test data, whereas corpus-level analyses were performed across the entire test set. Both approaches were applied to the top-performing model for each combination of REACH-VET risk tier and time window. A comparison to BoW interpretation approaches was deemed out of scope given extensive prior work in this area.

#### Note-Level Interpretation

Because the modeling framework employed gradient-boosted trees trained on fixed contextual embeddings averaged across all words in each note (*mean pooling*), gradient-based attribution methods were not applicable. Instead, note-level interpretability was achieved by combining Shapley Additive Explanations (SHAP) with layer activation mapping ^26–29^. SHAP analysis was used to identify the note-level embedding dimensions most influential in predicting suicide risk within the XGBoost model. For the most important embedding features, each word’s contextual representation was compared with the corresponding note-level average across those same dimensions. Inspired by the KeyBERT (keyword extraction for BERT) approach ^30,31^, the extent to which an individual word’s embedding diverged from the document average was calculated, indicating its relative influence on the suicide-related semantic language represented in the note-level embedding (**Figure 1C**). As a general interpretation, word importance scores were calculated at the note level and aggregated across randomly selected test-set notes. Privacy-preserving strategies were adopted for sharing note-level text and attribution maps (see **Supplementary Materials, “*Privacy-Preserving Generation of Representative Clinical Notes for Attribution Visualization*”**).

#### Corpus-Level Interpretation

At the corpus level, we examined semantic groupings of clinical notes to characterize broader themes captured by the top performing language models across each risk tier and prediction time window ^32^. To summarize and interpret patterns across all test-set notes, we applied BERTopic’s topic modeling methodology to SHAP-derived embeddings from the top-performing LLM for each stratum (**Figure 1D**). These SHAP-based embeddings act as supervised representations that emphasize suicide-relevant semantic dimensions beyond those present in the original pretrained embeddings. These note-level embeddings were first reduced to a lower-dimensional space using Uniform Manifold Approximation and Projection (UMAP) and subsequently clustered using Gaussian Mixture Models (a clustering algorithm) ^33–35^, with the optimal number of clusters selected via the Integrated Complete Likelihood criterion. For each resulting cluster, the LLM-interpretation framework identified uniquely associated words using class-based TF-IDF (cTF-IDF; a ‘bag-of-words’ approach commonly used to interpret LLM output in simpler terms) and calculated both the predicted proportion of suicide cases and the observed (true) proportion of suicide case notes. Finally, word rankings derived from cTF-IDF were compared across clusters to assess how the relative relevance of specific terms increased or decreased in association with higher cluster-level suicide proportions.

## Results

### Model Performance

For each of the nine combinations of suicide risk tier (low, moderate, high) and prediction window (<30, <90, <270 days), XGBoost models were trained using either bag-of-words (BoW) features or zero-shot (LLM embeddings, with and without the inclusion of structured patient characteristics. Patient-level performance estimates and 95% confidence intervals are reported in **Tables 2-3** and **Figure 2**.

**Figure 2:**
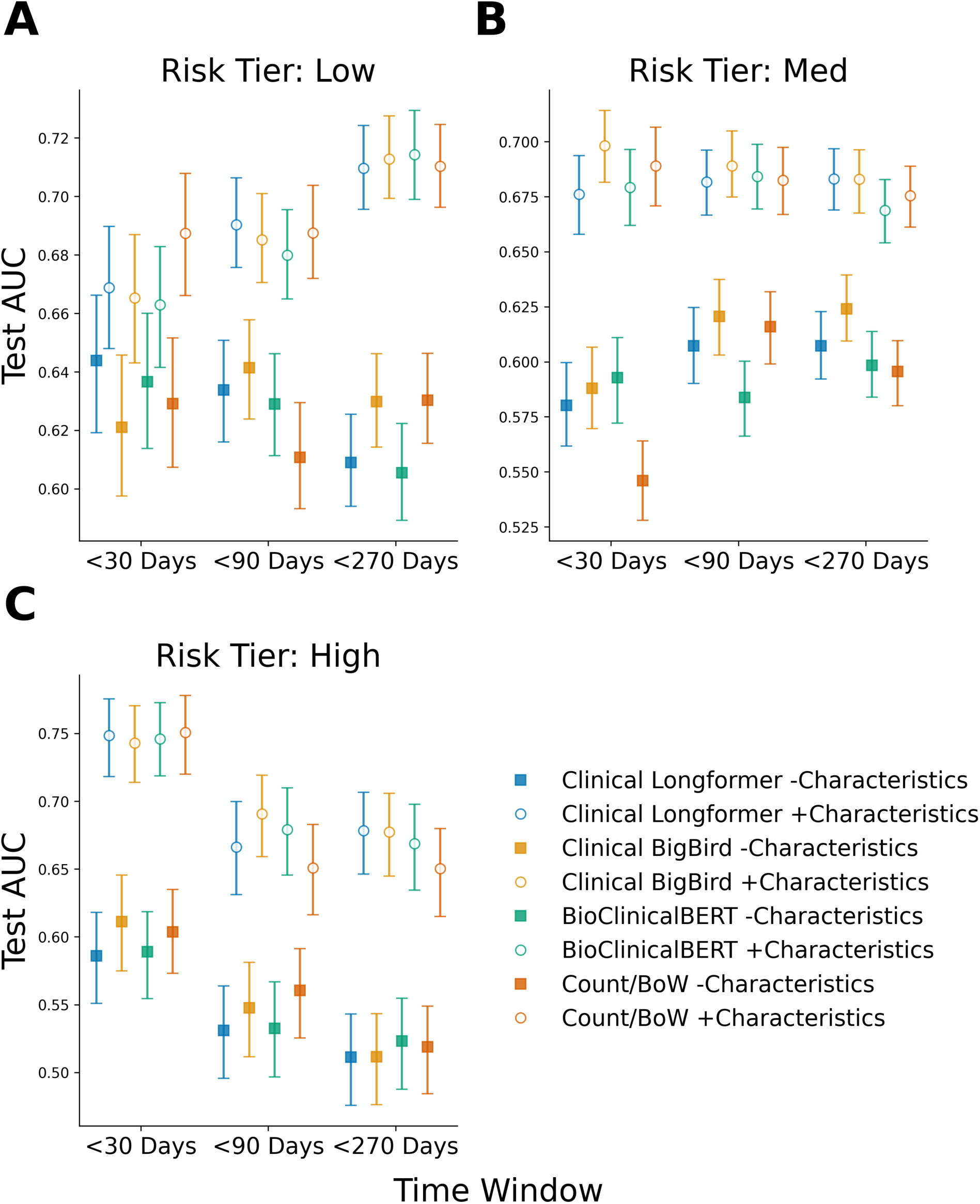
Model performance across risk tiers and prediction windows. Patient-level test-set area under the receiver operating characteristic curve (AUROC) for suicide risk prediction models stratified by REACH-VET risk tier: **A)** Low, **B)** Moderate/Medium, **C)** High, and prediction window (<30, <90, <270 days). Points represent mean AUROC values, and vertical bars denote 95% confidence intervals estimated via 1,000 bootstrap resamples. Models were trained using either text features alone or text combined with structured patient characteristics, and include pretrained large language model embeddings (Clinical Longformer, Clinical BigBird, BioClinicalBERT) and a bag-of-words (Count/BoW) baseline.

**Table 2.**
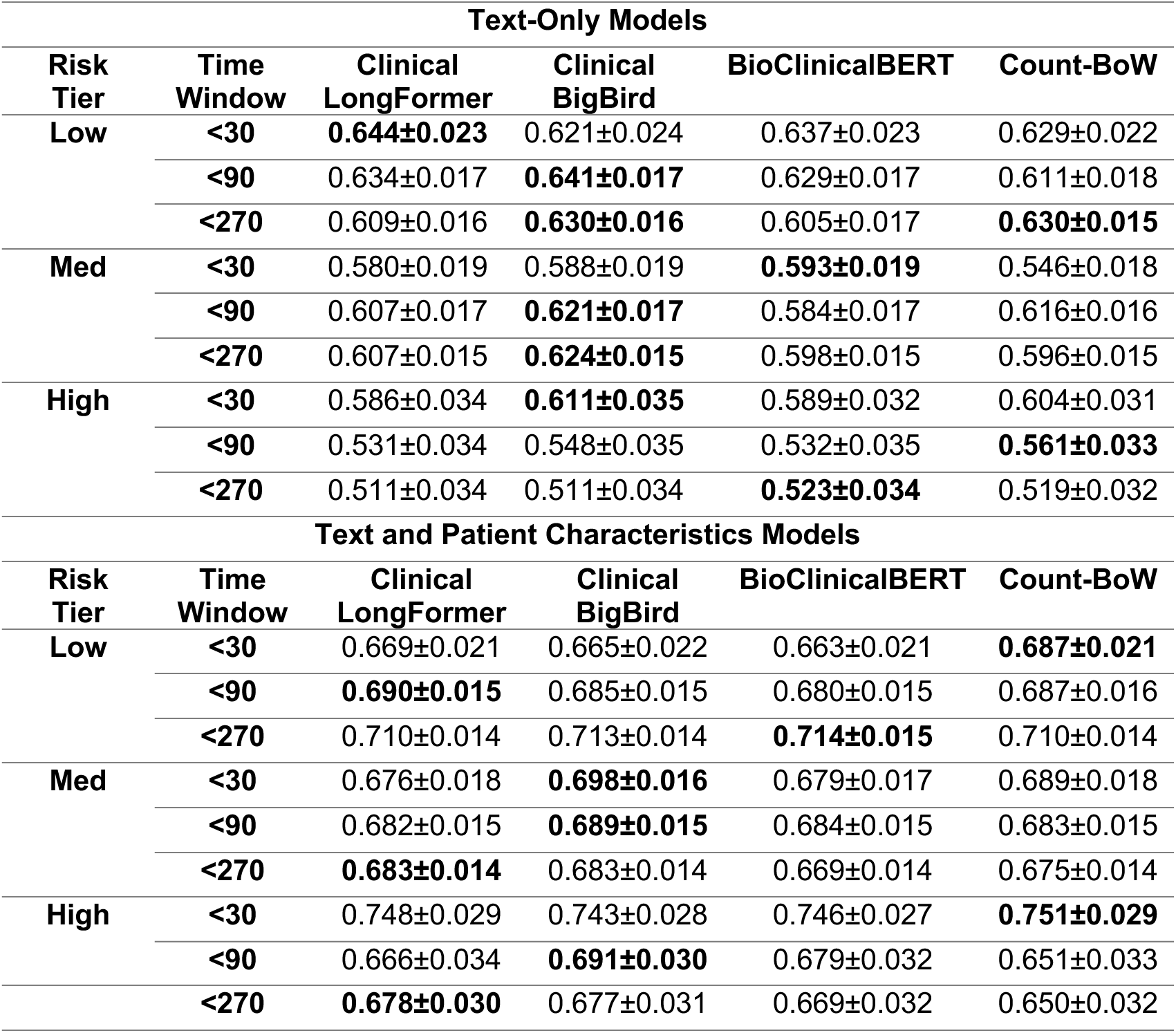
Model Performance Comparison. Top: Text-Only Performance. Patient-level area under the receiver operating characteristic curve (AUROC) for suicide risk prediction models trained using text features only. Models were trained separately within each combination of REACH-VET risk tier (low, moderate, high) and prediction window (<30, <90, <270 days). Results are shown for three pretrained large language model (LLM) embeddings (Clinical Longformer, Clinical BigBird, and BioClinicalBERT) and a bag-of-words (BoW) baseline. **Bottom: Text plus patient characteristics model performance.** Patient-level AUROC for suicide risk prediction models trained using text features combined with structured patient characteristics. Structured variables included demographics, clinical diagnoses, medication history, and healthcare utilization measures. Performance is reported across REACH-VET risk tiers and prediction windows for LLM-based embeddings and BoW representations. Values for both top and bottom tables represent patient-level test-set AUROC ± standard errors (used to estimate 95% confidence intervals), estimated via 1,000 bootstrap resamples with replacement.

#### Text-Only Models

When models were trained using text features alone (**Table 2**, **Figure 2**), LLM-based approaches demonstrated comparable or improved performance relative to BoW models in seven of nine risk tier–time window combinations. This advantage was most pronounced in the low- and moderate-risk tiers, where LLMs outperformed BoW approaches in five of six combinations by an average 4.3% (Range: 0.8%-8.6%) relative increase in AUROC for the top performing LLM compared to BoW, whereas gains in the high-risk tier were more modest (approximately 1% on average). While LLMs demonstrated strong performance at shorter time windows, BoW models occasionally matched or exceeded LLM performance at longer horizons, including the highest AUROC at <270 days (0.630).

Among the LLMs evaluated, BigBird achieving the highest or near-highest AUROC values across most strata. In the low-risk tier, Longformer yielded the top-performing LLM result at <30 days (0.644), while BigBird achieved the best LLM performance at <90 days (0.641) and matched the top LLM performance at <270 days (0.630). Similarly, in the moderate-risk tier, BigBird achieved the highest AUROC at <90 days (0.621) and <270 days (0.624), while BioClinicalBERT achieved the highest AUROC at <30 days (0.593). In the high-risk tier, BigBird achieved the highest LLM performance at <30 days (0.611), while BioClinicalBERT achieved the highest AUROC at <270 days (0.523).

#### Models Incorporating Patient Characteristics

The inclusion of structured patient characteristics led to substantial performance improvements across all modeling approaches (**Table 2**), with AUROC gains of approximately 15.7% (Range: 6.7% - 29.6%) relative to text-only models. Pretrained LLM embeddings continued to demonstrate competitive or superior performance relative to BoW representations in seven of nine risk tier–time window combinations.

Within this multimodal setting, BigBird and Longformer again emerged as the most performant LLMs, together achieving the highest AUROC among LLMs in eight of nine strata. Notably, Longformer achieved the best LLM performance in the low-risk tier at <90 days (0.690), while BioClinicalBERT achieved the highest LLM performance at <270 days (0.714). BigBird achieved the best overall LLM performance in the high-risk tier at <90 days (0.691). While BoW models achieved the highest AUROC in select settings, notably in the high-risk tier at <30 days (0.751), BigBird’s performance was generally comparable and often superior to other LLM variants.

#### Temporal Trends by Risk Tier

Across text representations, temporal trends in model performance were observed by risk tier. In the low-risk tier, performance improved with increasing prediction window length, with AUROC values rising from approximately 0.64– 0.67 at <30 days to 0.71-0.72 at <270 days when patient characteristics were included. In the high-risk tier, performance consistently peaked at <30 days and declined at longer time windows, with AUROC values decreasing from 0.586–0.611 to 0.511–0.523 for text-only models and from 0.743–0.751 to 0.669–0.678 when patient characteristics were included (**Table 2**).

### Model Interpretation

**Figure 3:**
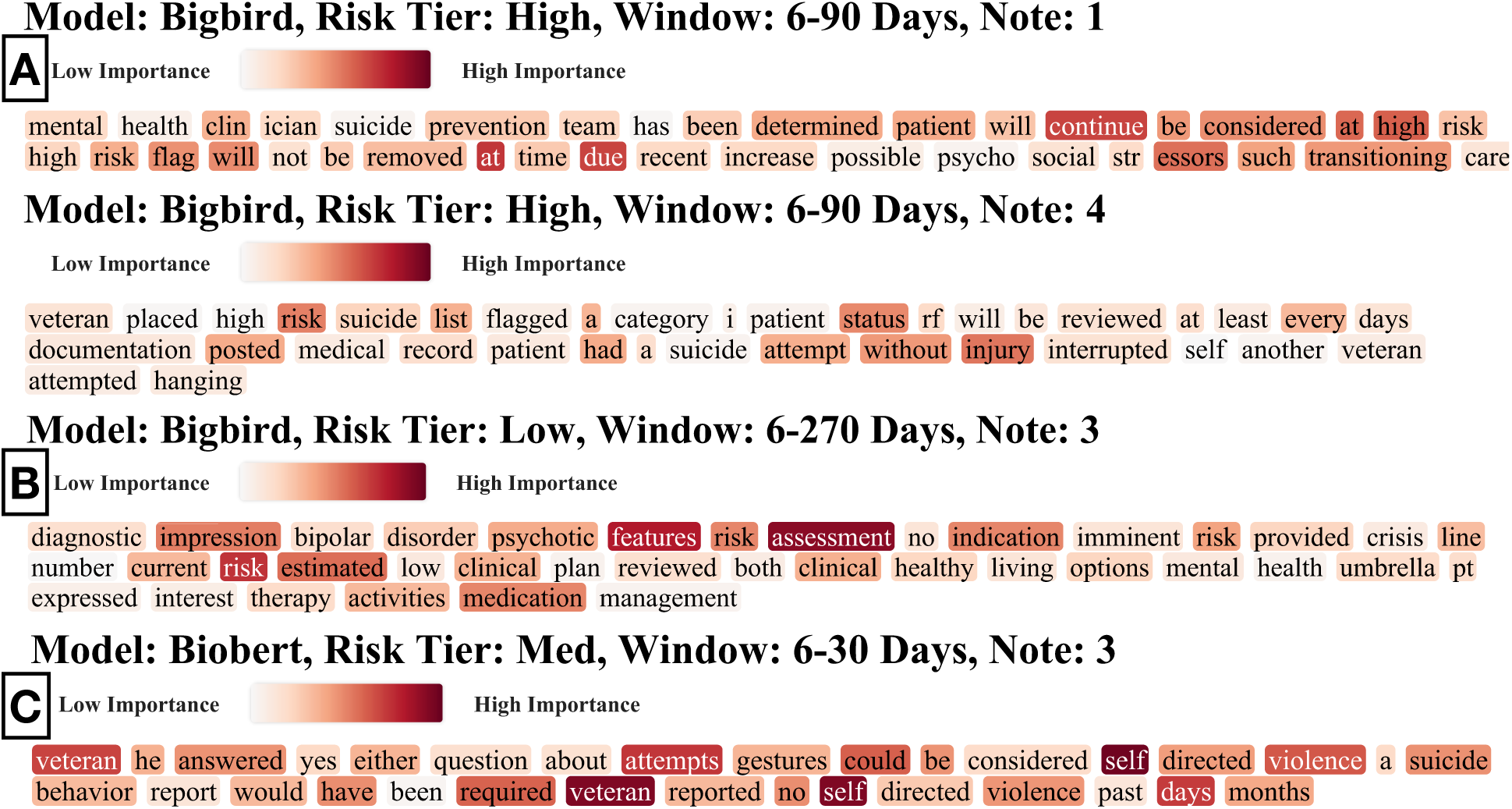
Representative note-level word importance maps for: **A)** Clinical BigBird (high risk, 6–90 days), **B)** Clinical BigBird (low risk, 6–270 days), and **C)** BioClinicalBERT (moderate risk, 6-30 days). Color intensity denotes relative contribution of individual words to model predictions. Clinical synopsis text was generated from original clinical reports using a locally deployed, privacy-preserving large language model after iterative note-level de-identification using named entity recognition followed by manual review. The rewritten text preserved clinically meaningful structure while removing personally identifiable information and retaining selected high-importance tokens. The resulting text was processed through the LLM embedding model, XGBoost classification, and SHAP workflows to compute token-level feature attribution scores quantifying the contribution of individual words to the predicted suicide risk. Words highlighted in red indicate greater relative importance in the model’s prediction. For presentation purposes, the final generated text and attribution maps were further subset to prioritize less patient-specific language and improve clarity of the visualization.

**Table 3:** High-importance words aggregated across randomly selected test-set clinical notes for each language model. Words were ranked by their relative importance scores across individual reports, which were then averaged across all reports and finally reweighted using the inverse-document frequency (λ=0.001).

| Model | Ranked High-Importance Words |
| --- | --- |
| Clinical Longformer | harm, minute, done, thought, suicide, see, continue, bed, veteran, attempt, patient, date, every, assessment, time, given, plan, report, diagnosis, being, history, problem, any, suicidal, risk, alert, change, unit, concern, night, name, medication, place, medical, restraint, self, service, care, level, current |
| BioClinicalBERT | blood, shift, minute, assessment, assistance, mental, assist, name, family, within, symptom, day, treatment, monitor, note, being, medication, fall, any, high, call, hygiene, safety, intervention, none, evaluation, status, |
|  | patient, care, all, continue, once, suicide, psychiatric, offer, signature, self, report, environment, effect |
| **Clinical BigBird** | call, name, note, veteran, time, pain, needed, patient, score, hour, self, need, unit, check, document, any, phone, report, medication, day, care, date, given, continue, reported, contact, diagnosis, other, bed, risk, assessment, blood, past, plan, following, sign, state, alert, admitted, hospital |

#### Note-Level Interpretation

Across notes, high-importance words were concentrated within a relatively small subset of words, with the most influential terms consistently reflecting clinically salient themes (**Table 3**). These included explicit references to suicide attempts and ideation, indicators of outreach or support engagement, expressions of crisis or distress, and contextual factors such as pain and sleep disturbance. In addition, models frequently assigned elevated importance to terms related to clinical risk flagging and assessment. Representative examples of note-level word importance visualizations are shown in **Figure 3** and **Supplementary Figures 1-9**.

#### Corpus-Level Interpretation

Across topics (document clusters), semantic content varied systematically with suicide risk (**Supplementary Figure 10; Supplementary Tables 1-2**). In addition, the relative importance of specific words within clusters shifted as suicide risk increased (**Supplementary Figures 11-12**). For example, in the high-risk tier (<90 days), the BigBird model identified that treatment-related terms and reports of pain had diminishing prominence, while references to suicide planning became increasingly prominent. A similar pattern was observed in the BioClinicalBERT model within the moderate-risk tier (<90 days), where language related to ongoing care and treatment decreased in relevance as cluster-level suicide proportions increased, whereas explicit mentions of suicide planning became more salient.

When examined across topics for the top LLM models identified within each risk tier–time window combination, clear thematic patterns emerged. In the low-risk tier, topics were dominated by routine medical care and administrative documentation. Across all time windows, recurrent high-weight terms reflected medication management and procedural language (e.g., “one tablet”, “refill”, “mg”, “expiration”), physical health and symptom monitoring (e.g., “pressure”, “pain”, “sleep”, “wound”, “skin”), and care logistics (e.g., “clinic”, “provider”, “contact”, “supply”). In the moderate-risk tier, explicit suicide assessment and planning language emerged more consistently across topics. Shorter time windows were characterized by frequent references to suicide and planning (e.g., “suicide”, “plan”, “critical”, “risk”), alongside ongoing medical monitoring and treatment (e.g., “medication”, “mg”, “tablet”, “assessment”). At longer horizons, topics increasingly emphasized functional status and chronic impairment (e.g., “walk”, “independent”, “pain”, “physical”). In the high-risk tier, topics across all time windows prominently featured safety monitoring, staff involvement, and explicit suicide-related language. Across multiple clusters, high-importance terms included references to suicide and self-harm (e.g., “suicide”, “self”, “thought”, “plan”), safety and supervision (e.g., “staff support”, “safety”, “monitor”, “risk”), and functional performance (e.g., “activity”, “mobility”). At longer prediction windows, these themes were accompanied by increased emphasis on care escalation and documentation (e.g., “intervention”, “ordered”, “discharge”, “goal”), as well as mood-related language (e.g., “mood”, “mental”).

## Discussion

There is growing interest in developing suicide risk prediction models that leverage unstructured clinical narratives to complement existing approaches based on structured EHR data, such as REACH-VET, within veteran populations. Prior work has demonstrated the utility of NLP methods for identifying psychosocial and linguistic patterns associated with suicide risk and informing monitoring programs ^36–40^. However, most existing approaches rely on count-based or bag-of-words representations that disregard linguistic context ^41,42^. In contrast, the findings presented here represent one of the first systematic efforts to evaluate LLM–based representations for suicide risk prediction in a large veteran cohort. By capturing contextual and semantic relationships within clinical text, LLMs offer a more nuanced understanding of clinical narratives than traditional lexical methods.

This study establishes an initial baseline for LLM-based suicide risk prediction using several pretrained architectures that have been previously explored in clinical text analysis. Importantly, these models were applied in an off-the-shelf, zero-shot setting without additional fine-tuning. Even under this conservative framework, deep learning-based representations demonstrated superior or comparable performance to bag-of-words approaches in several clinically relevant contexts. While further improvements are likely achievable through domain-specific fine-tuning or temporal modeling, such extensions were beyond the scope of the present work and intentionally deferred to establish a clear baseline for comparison.

Several key patterns emerged from our results. Among individuals in lower suicide risk tiers, predictive performance improved as the predictive time window lengthened, suggesting that longer-term clinical documentation provides additional predictive information for distinguishing suicide risk among lower-risk patients. In contrast, within the high-risk tier, performance was highest at shorter time windows and declined at longer horizons, regardless of text representation. This pattern indicates that short-term clinical language may be particularly informative for identifying imminent suicide risk among high-risk individuals, whereas longer-term documentation may dilute or obscure acute risk signals.

These performance trends were concordant with findings from corpus-level topic modeling. In lower-risk groups, topics were dominated by routine management, procedural documentation, and care logistics, consistent with longitudinal patterns of ongoing medical care. In contrast, among high-risk patients, particularly closer to the time of death, clinical language more frequently included explicit references to suicidal ideation, planning, or attempts. At longer horizons, even within higher-risk tiers, topics increasingly reflected chronic medical issues and functional impairments rather than acute suicide-related language. Further, our findings suggested that corpus-level groupings derived from LLM embeddings can meaningfully track suicide risk across document themes, illustrating how the relevance and usage of specific terms shift as risk increases.

Interestingly, terms that might initially appear unrelated to suicide risk, such as references to skin, bowel, or other somatic concerns, emerged as prominent within certain topics. These findings may reflect the role of chronic medical conditions that are known to co-occur with elevated suicide risk (e.g., cancer, inflammatory bowel disease), as well as the downstream effects of acute medical interventions (e.g., in-patient care prior to suicide attempt or complications following prior suicide attempts) or withdrawal-related symptoms documented in clinical notes ^43–45^. Such language may therefore serve as indirect markers of vulnerability rather than spurious signals.

These findings affirm noted service utilization trends, with patients in lower suicide risk tiers tending to use less mental health services and be less disclosive about suicidality and patients in high suicide risk tiers having the opposite tendencies ^46^. In contrast to established risk metrics, such as REACH-VET, which highly weight mental health usage and prior suicide attempts in suicide risk modeling, this methodology offers heightened ability to identify and monitor these discrete populations’ risk trajectories. This innovation offers to capture greater predictive accuracy and clinical nuance among low-risk patients, a population who represents the majority of VA suicide decedents, yet are understudied and underserved in clinical care.

Several limitations of this study warrant consideration and motivate directions for future work. First, based on our findings, clinical notes drawn from lower suicide risk tiers and longer prediction horizons are likely to be less directly associated with suicide-related outcomes. We aimed to capture longitudinal care trajectories across all provider types, including primary care, given their role in clinical decision-making and referral pathways. Nonetheless, not all clinical notes, whether mental health-related or not, are equally relevant for suicide risk assessment. Many notes are procedural or administrative in nature and may not reflect evaluative content or patient response to follow-up, which could dilute the predictiveness of the text.

Second, the current modeling framework does not explicitly account for the temporal ordering of notes within individual patients. The sequence in which notes are written may carry important contextual information, as the interpretation of a given note may depend on preceding documentation. This temporal dependency is not captured by the present approach, which aggregates notes within predefined time windows. Future work should explore temporal modeling strategies that incorporate both content and timing, enabling identification of key inflection points for intervention and more precise filtering of clinically relevant documentation beyond coarse time window categorization ^47–49^. Implementing such approaches may be particularly challenging in lower-risk populations and at longer time horizons, where the proportion of less relevant documentation is likely higher. In addition, baseline risk estimates such as REACH-VET used to group patients into risk strata may be less stable in settings with sparser or more fragmented records.

Third, while this study evaluated several established LLM architectures, rapid advances in deep learning have produced newer models pretrained on larger and more varied corpora that may offer improved performance ^50–52^. The models used here were selected based on feasibility within a highly privacy-preserving computational environment. As such, these results should be interpreted as establishing a baseline rather than reflecting the maximal achievable performance. Moreover, the clinical notes analyzed in this study were drawn from a 2017–2018 corpus and may not capture subsequent changes in clinical workflows, documentation practices, or evolving language related to suicidality. While this historical data is sufficient for comparative evaluation of predictive performance, deployment of such models would require recalibration using more contemporary clinical data. Finally, this work does not explicitly address provider misspellings or idiosyncratic documentation practices ^53,54^, which may affect tokenization and downstream representation of clinical text. While improved tokenization or domain-adapted fine-tuning could mitigate some of these issues, such refinements were beyond the scope of the current study and may have influenced both predictive performance and interpretability.

There are substantial opportunities for further exploration building on the present work. Early NLP efforts in suicide risk modeling focused on well-curated semantic lexicons designed to quantify predefined psychosocial constructs by counting matched terms. More recently, growing interest has shifted toward generative AI approaches that use prompt-based strategies to elicit higher-level psychosocial attributes directly from clinical text ^55–57^. Such approaches allow models to infer constructs such as treatment modality, suicide-related language, or clinical engagement without requiring explicit specification of search terms, potentially enabling more flexible and context-sensitive feature extraction. Importantly, this paradigm may also help disentangle provider-specific documentation styles, such as linguistic markers of therapeutic alliance, that can otherwise confound interpretations of patient risk trajectories. However, implementation may require substantial task supervision, careful prompt design, and validation to ensure reliable interpretation and extraction.

In doing this research, we observed that integrating demographic and clinical characteristics substantially improved predictive performance. Future work could further incorporate structured patient information into end-to-end deep learning architectures, enabling more nuanced modeling of interactions between language and patient characteristics. Such integration may improve performance within clinically heterogeneous subgroups and facilitate identification of modifying risk factors across sub-cohorts. Ultimately, translation of these methods into clinical practice will require additional fine-tuning, prospective validation, and careful collaboration with clinical and operational stakeholders to ensure interpretability, fairness, and safe implementation.

## Data Availability

The data that support the findings of this study were obtained from the U.S. Department of Veterans Affairs (VA) Corporate Data Warehouse and the VA Department of Defense Mortality Data Repository. These data contain sensitive patient information and are not publicly available. Access to VA data is subject to federal privacy regulations, institutional approvals, and data use agreements. Qualified researchers may request access through the VA Informatics and Computing Infrastructure (VINCI) or other VA data governance mechanisms. Analytic code may be made available from the corresponding author upon reasonable request.

## Acknowledgements

This work is supported by Department of Defense grant PR220927 / HT9425-23-1-0267 to JL, ML, JG, BS, NIH P30CA023108 support for JL, and VA Clinical Science Research Career Development Award (CX002630) to ML. Our analysis was completed using a project specific VA research server and Prospect, a designated Amazon Web Service VA Enterprise Cloud enclave.

