## Supplementary Materials for "Comparative Evaluation of Pretrained Large Language Models for Suicide Risk Prediction from Clinical Notes in U.S. Veterans"

#### Supplementary Method: Privacy-Preserving Generation of Representative Clinical Notes for Attribution Visualization

Because individual clinical notes cannot be shared publicly due to institutional review board (IRB) restrictions, even when de-identified, as reporting must occur at an aggregate level (as done above), the original text used to derive word importance scores could not be presented directly in this manuscript. To illustrate note-level attribution patterns while preserving confidentiality, we developed an alternative visualization strategy. Representative clinical synopsis text was generated from original notes using a locally deployed, privacy-preserving large language model. Prior to generation, notes underwent iterative de-identification using automated named entity recognition followed by manual review to remove personally identifiable information. The generated text was designed to maintain high semantic fidelity to the original clinical content while preserving clinically meaningful structure and selected high-importance tokens identified in the original attribution analyses. Study investigators reviewed generated synopses to ensure alignment with the source content and additional removal of identifying details (e.g., mention of specific/regional VA medical center name). The resulting text was then processed through the same embedding model as the original text, XGBoost classification pipeline, and SHAP feature attribution workflow to compute token-level importance scores reflecting the contribution of individual words to predicted suicide risk. Attributions were examined again for visual alignment with the original attributions and preservation of important suicide-related terminology. For visualization purposes, text within selected examples were further subset to prioritize less patient-specific language and improve interpretability of the attribution maps.

### Model: Longformer, Risk Tier: Low, Window: 6-30 Days, Note: 1

Low Importance High Importance

had su icidal ide ation plan shoot himself approximately years ago no other incidents history depression su icidal ide ation plan years ago elderly male c a uc ian likely cognitive impairment protective factors good mood no si past years good ocial support coping skills future oriented thinking based risk protective factors patient low risk suicide self harm

### Model: Longformer, Risk Tier: Low, Window: 6-30 Days, Note: 2

Low Importance High Importance

wife she stated patient has mood swing does not feel she danger we discussed option call she has her safety also gave crisis line number

### Model: Longformer, Risk Tier: Low, Window: 6-30 Days, Note: 3

Low Importance High Importance

self identified anxiety he last cut himself days ago area band aged veteran said he feels like giving right now he has been trying cope his own he can no longer do he reached support

### Model: Longformer, Risk Tier: Low, Window: 6-30 Days, Note: 4

Low Importance High Importance

he denies having a plan carry suicide his own saying i just wanted learn more about since i about tv i dont want go through what my went through not at my age plan discussion p cp crisis nurse intervention si veteran will wait discussion crisis intervention

**Supplementary Figure 1: Representative note-level word importance maps for Clinical LongFormer (low risk, 6–30 days).** Clinical synopsis text was generated from original clinical reports using a locally deployed, privacy-preserving large language model after iterative note-level de-identification using named entity recognition followed by manual review. The rewritten text preserved clinically meaningful structure while removing personally identifiable information and retaining selected high-importance tokens. The resulting text was processed through the LLM embedding model, XGBoost classification, and SHAP workflows to compute token-level feature attribution scores quantifying the contribution of individual words to the predicted suicide risk. Words highlighted in red indicate greater relative importance in the model's prediction. For presentation purposes, the final generated text and attribution maps were further subset to prioritize less patient-specific language and improve clarity of the visualization.

### Model: Longformer, Risk Tier: Low, Window: 6-90 Days, Note: 1

Low Importance High Importance

counted narc otic pills did not exceed usual doses they were sorts a day incident patient has many reasons live denies suicidal risk

### Model: Longformer, Risk Tier: Low, Window: 6-90 Days, Note: 2

Low Importance High Importance

reported depression a long time could not give more details beyond many thoughts he expressed worry about life denied suicidal thought plan intent he assisted learning a grounding technique directed exercise he found helpful will try at home he given veteran crisis line his psychiatrist will refer him a psychologist

### Model: Longformer, Risk Tier: Low, Window: 6-90 Days, Note: 3

Low Importance High Importance

he den ies active plan intent kill himself others no current indication a safety welfare evaluation

**Supplementary Figure 2: Representative note-level word importance maps for Clinical LongFormer (low risk, 6–90 days).** Clinical synopsis text was generated from original clinical reports using a locally deployed, privacy-preserving large language model after iterative note-level de-identification using named entity recognition followed by manual review. The rewritten text preserved clinically meaningful structure while removing personally identifiable information and retaining selected high-importance tokens. The resulting text was processed through the LLM embedding model, XGBoost classification, and SHAP workflows to compute token-level feature attribution scores quantifying the contribution of individual words to the predicted suicide risk. Words highlighted in red indicate greater relative importance in the model's prediction. For presentation purposes, the final generated text and attribution maps were further subset to prioritize less patient-specific language and improve clarity of the visualization.

#### Model: Bigbird, Risk Tier: Low, Window: 6-270 Days, Note: 1

Low Importance High Importance

social occupational oc impairment attention vulnerability around triggering patient denied suicidal hom icidal ide ation plan intent goal establish rapport will involve mixing history

#### Model: Bigbird, Risk Tier: Low, Window: 6-270 Days, Note: 2

Low Importance High Importance

a self care session clinical issue collaborative decision focus now issues immediate future patient will consult community psychologist psychiatrist medication current issue patient denied suicidal hom icidal ide ation plan intent goal establish rapport

**Supplementary Figure 3: Representative note-level word importance maps for Clinical BigBird (low risk, 6–270 days).** Clinical synopsis text was generated from original clinical reports using a locally deployed, privacy-preserving large language model after iterative note-level de-identification using named entity recognition followed by manual review. The rewritten text preserved clinically meaningful structure while removing personally identifiable information and retaining selected high-importance tokens. The resulting text was processed through the LLM embedding model, XGBoost classification, and SHAP workflows to compute token-level feature attribution scores quantifying the contribution of individual words to the predicted suicide risk. Words highlighted in red indicate greater relative importance in the model's prediction. For presentation purposes, the final generated text and attribution maps were further subset to prioritize less patient-specific language and improve clarity of the visualization.

### Model: Biobert, Risk Tier: Med, Window: 6-30 Days, Note: 1

Low Importance High Importance

morning hung phone at approximately pm throughout call police were informed veteran s risk harm himself dr notified at approximately pm he made attemts contact without success signed

### Model: Biobert, Risk Tier: Med, Window: 6-30 Days, Note: 2

Low Importance High Importance

he denied suicidal ideation plan intent does not believe suicide would help he does not want provider call police he plans get through night

**Supplementary Figure 4: Representative note-level word importance maps for BioClinicalBERT (moderate risk, 6–30 days).** Clinical synopsis text was generated from original clinical reports using a locally deployed, privacy-preserving large language model after iterative note-level de-identification using named entity recognition followed by manual review. The rewritten text preserved clinically meaningful structure while removing personally identifiable information and retaining selected high-importance tokens. The resulting text was processed through the LLM embedding model, XGBoost classification, and SHAP workflows to compute token-level feature attribution scores quantifying the contribution of individual words to the predicted suicide risk. Words highlighted in red indicate greater relative importance in the model's prediction. For presentation purposes, the final generated text and attribution maps were further subset to prioritize less patient-specific language and improve clarity of the visualization.

**Model: Bigbird, Risk Tier: Med, Window: 6-90 Days, Note: 1**

Low Importance High Importance

suicide prevention coordinator able get a hold pt am pt ins instant he not suicidal had no plan harm self pt also stated his intent

**Model: Bigbird, Risk Tier: Med, Window: 6-90 Days, Note: 2**

Low Importance High Importance

shift nursing note team obtained vital signs current sleep pattern at bed side patient apparently asleep bed maintained calm demeanor denied suicidal thoughts

**Supplementary Figure 5: Representative note-level word importance maps for Clinical BigBird (moderate risk, 6–90 days).** Clinical synopsis text was generated from original clinical reports using a locally deployed, privacy-preserving large language model after iterative note-level de-identification using named entity recognition followed by manual review. The rewritten text preserved clinically meaningful structure while removing personally identifiable information and retaining selected high-importance tokens. The resulting text was processed through the LLM embedding model, XGBoost classification, and SHAP workflows to compute token-level feature attribution scores quantifying the contribution of individual words to the predicted suicide risk. Words highlighted in red indicate greater relative importance in the model's prediction. For presentation purposes, the final generated text and attribution maps were further subset to prioritize less patient-specific language and improve clarity of the visualization.

### Model: Longformer, Risk Tier: Med, Window: 6-270 Days, Note: 1

Low Importance High Importance

ve ter presented alert oriented angry mood con gru ent affect organized goal oriented thought clear speech he discussed he ha had anxiety his whole life became depressed

### Model: Longformer, Risk Tier: Med, Window: 6-270 Days, Note: 2

Low Importance High Importance

medical condition occurs dangerous situations d ependence represents a more severe state characterized failed attempts ab st ain control use physiological dependence ind icated increased tolerance withdrawal symptoms such n ause a trem ors mor use stops common mental health disorders among those substance dependence include mood anxiety disorders a higher proportion individuals severe mental illness including schizophrenia izo sch izo aff ective disorder also have co occ urring substance use problems these disorders can

**Supplementary Figure 6: Representative note-level word importance maps for Clinical LongFormer (moderate risk, 6–270 days).** Clinical synopsis text was generated from original clinical reports using a locally deployed, privacy-preserving large language model after iterative note-level de-identification using named entity recognition followed by manual review. The rewritten text preserved clinically meaningful structure while removing personally identifiable information and retaining selected high-importance tokens. The resulting text was processed through the LLM embedding model, XGBoost classification, and SHAP workflows to compute token-level feature attribution scores quantifying the contribution of individual words to the predicted suicide risk. Words highlighted in red indicate greater relative importance in the model's prediction. For presentation purposes, the final generated text and attribution maps were further subset to prioritize less patient-specific language and improve clarity of the visualization.

#### Model: Bigbird, Risk Tier: High, Window: 6-30 Days, Note: 1

**Low Importance** **High Importance**

depression suicidal ide ation history present illness year old gentleman a diagnosis major recurrent depressive disorder patient requesting voluntary admission psychiatric facility earlier

#### Model: Bigbird, Risk Tier: High, Window: 6-30 Days, Note: 2

**Low Importance** **High Importance**

expressed having a hard time dealing personal distress stating fatigue memories explosion went bad veteran reported purchasing a gun days ago had been pulling trigger bullet chamber each day past days

#### Model: Bigbird, Risk Tier: High, Window: 6-30 Days, Note: 3

**Low Importance** **High Importance**

has been seen both myself other staff demonstrate no impuls ivity depression indication elevated safety concern would have contra ind icated return ideally

**Supplementary Figure 7: Representative note-level word importance maps for Clinical BigBird (high risk, 6–30 days).** Clinical synopsis text was generated from original clinical reports using a locally deployed, privacy-preserving large language model after iterative note-level de-identification using named entity recognition followed by manual review. The rewritten text preserved clinically meaningful structure while removing personally identifiable information and retaining selected high-importance tokens. The resulting text was processed through the LLM embedding model, XGBoost classification, and SHAP workflows to compute token-level feature attribution scores quantifying the contribution of individual words to the predicted suicide risk. Words highlighted in red indicate greater relative importance in the model's prediction. For presentation purposes, the final generated text and attribution maps were further subset to prioritize less patient-specific language and improve clarity of the visualization.

### Model: Bigbird, Risk Tier: High, Window: 6-90 Days, Note: 1

**Low Importance** **High Importance**

veteran s high risk status recently reviewed suicide prevention staff veteran s m h provider veteran receiving service another va mc no longer followed at mc high risk flag suicide being transferred due reason veteran now followed at mc suicidal

### Model: Bigbird, Risk Tier: High, Window: 6-90 Days, Note: 2

**Low Importance** **High Importance**

suicide prevention team received notification a veteran s self reported suicide attempt occurred veteran overd osed sleeping medication taking several tablets sitting a bench outside taking medication veteran contacted v crisis line v line initiated a rescue

### Model: Bigbird, Risk Tier: High, Window: 6-90 Days, Note: 3

**Low Importance** **High Importance**

patient reports ongoing thoughts hurt himself no concrete plan year old male admitted bipolar depression veteran went er at stated he having suicidal thoughts overdose hang himself

### Model: Bigbird, Risk Tier: High, Window: 6-90 Days, Note: 4

**Low Importance** **High Importance**

high risk suicide p rf designation has been added veteran s chart a category i patient record flag high risk suicide applied status p rf will be reviewed at least every day documentation posted medical record a comment notes a suicide attempt without injury

**Supplementary Figure 8: Representative note-level word importance maps for Clinical BigBird (high risk, 6–90 days).** Clinical synopsis text was generated from original clinical reports using a locally deployed, privacy-preserving large language model after iterative note-level de-identification using named entity recognition followed by manual review. The rewritten text preserved clinically meaningful structure while removing personally identifiable information and retaining selected high-importance tokens. The resulting text was processed through the LLM embedding model, XGBoost classification, and SHAP workflows to compute token-level feature attribution scores quantifying the contribution of individual words to the predicted suicide risk. Words highlighted in red indicate greater relative importance in the model's prediction. For presentation purposes, the final generated text and attribution maps were further subset to prioritize less patient-specific language and improve clarity of the visualization.

### Model: Biobert, Risk Tier: High, Window: 6-270 Days, Note: 1

**Low Importance** **High Importance**

voluntary patient reported suicidal thoughts impulses continued substance use interfere functioning ptsd symptoms major depression isolation alcohol abuse angry

### Model: Biobert, Risk Tier: High, Window: 6-270 Days, Note: 2

**Low Importance** **High Importance**

interferes function exacerbation symptoms isolation alcohol abuse angry burst guilt shame hedonia associated diagnosis major depression referral

**Supplementary Figure 9: Representative note-level word importance maps for Clinical BioClinicalBERT (high risk, 6–270 days).** Clinical synopsis text was generated from original clinical reports using a locally deployed, privacy-preserving large language model after iterative note-level de-identification using named entity recognition followed by manual review. The rewritten text preserved clinically meaningful structure while removing personally identifiable information and retaining selected high-importance tokens. The resulting text was processed through the LLM embedding model, XGBoost classification, and SHAP workflows to compute token-level feature attribution scores quantifying the contribution of individual words to the predicted suicide risk. Words highlighted in red indicate greater relative importance in the model's prediction. For presentation purposes, the final generated text and attribution maps were further subset to prioritize less patient-specific language and improve clarity of the visualization.

**Supplementary Table 1: Text-Only Topic Word Importances (cTF-IDF),** with separate topic modeling for reports stratified based on top performing LLM for risk tier and predictive time window, for models which did not integrate patient characteristics

| Risk Tier | Time Window | Model | Cluster | Suicide Risk Proportion | Top Words |
|---|---|---|---|---|---|
| **Low** | 30 | LongFormer | 1 | 0.23 | active, record, moisture, daily, unit, community, type, every, bowel movement, test |
| | | | 2 | 0.23 | alternate, sleep, service, group, visit, last, th, mouth active, need, twice |
| | | | 3 | 0.18 | active, treatment, mouth every, imaging, use, medical, set up, mouth active, every day, visit |
| | | | 4 | 0.22 | active, mgdl, mg, choice, office, do, test, eligibility, month, needed |
| | | | 5 | 0.22 | active, screening, month, daily, warfarin, do, unit ref, home, bowel, unit |
| | | | 6 | 0.44 | eligibility, medication, daily, day, screen, every day, collection, mg, condition, total |
| | | | 7 | 0.26 | active, screening, one tablet, day, unit, eval, urine, pt, bowel movement, test |
| | | | 8 | 0.28 | all, th, result unit, goal, up, moisture, specimen, often, how often, warfarin |
| | | | 9 | 0.42 | active, sleep, primary, lab, resident, how, set up, medication, call, sig |
| | | | 10 | 0.23 | range, community care, service, did, choice, dose, self, cm, screening, authorization |
| | 90 | BigBird | 1 | 0.22 | left, all, scanned, mental, every, remaining, session, group, care, pain |
| | | | 2 | 0.26 | plan, all, one tablet, left, need, day, procedure, daily, but, last |
| | | | 3 | 0.2 | tablet, active, service, needed, session, blood, suicide, remaining, every, scanned |
| | | | 4 | 0.19 | right, all, left, mg, document, goal, report, last, care, needed |
| | | | 5 | 0.19 | remaining, about, mouth, needed, every day, last, procedure, group, call, need |

| | | | | | |
|---|---|---|---|---|---|
| | | | 6 | 0.2 | right, about, day, medication, plan, contact, report, document, change, one |
| | | | 7 | 0.18 | right, about, need, goal, mg, change, remaining, filled, cm, one tablet |
| | | | 8 | 0.27 | assessment, medication, mouth every, mental, it, session, every, day, blood, one |
| | | | 9 | 0.16 | reviewed, about, left, need, it, contact, right, medication, cm, one |
| | | | 10 | 0.19 | remaining, about, one, provider, mouth every, day supply, refill, may, every day, mental |
| | 270 | | 1 | 0.18 | active, problem, procedure, report, per, daily, mg, day, given, medication |
| | | | 2 | 0.15 | active, pressure, past, one, pt, daily, every, day, medication, left |
| | | | 3 | 0.4 | an, fall, past, pressure, one tablet, cm, right, assessment, per, pt |
| | | | 4 | 0.22 | active, given, pain, tablet, mouth, needed, plan, care, per, doe |
| | | | 5 | 0.29 | call, per, one tablet, right, day, tablet, past, blood, up, mg |
| | | | 6 | 0.16 | active, one tablet, plan, skin, problem, every, procedure, daily, past, last |
| | | | 7 | 0.19 | active, one, procedure, use, fall, daily, plan, call, mg, one tablet |
| **Med** | 30 | BioBERT | 1 | 0.24 | plan care, mg, tablet, call, home, date, assessment, icd, mouth, prn |
| | | | 2 | 0.19 | required electronically, plan, transfer, ability, needed, education, day, pt, change, per |
| | | | 3 | 0.19 | scanned document, pressure, treatment, ability, prn, goal, click, required, assessment, filed |
| | | | 4 | 0.18 | need, functional, up, all, fall, document, change, overall, home, procedure |
| | | | 5 | 0.25 | can, call, pain, per, every, phone, goal, change, place, pressure |
| | | | 6 | 0.2 | respiratory, reassessment, scanned document, every day, per, every, continue, place, mood, reported |
| | | | 7 | 0.26 | reported, required electronically, clear, respiratory, place, goal, date, plan care, medication, rest |

| | | | | | |
|---|---|---|---|---|---|
| | | | 8 | 0.22 | without, mg, required electronically, ml, information, call, able, electronically filed, good, functional |
| | | | 9 | 0.2 | pressure, prn, tool, able, click, every, activity, overall, filed, ml |
| | | | 10 | 0.22 | without, jul, risk, information, overall, icd, able, plan, continue monitor, contact |
| | 90 | BigBird | 1 | 0.19 | active, refill, use, medication, mg, last, contact, sig, date, care |
| | | | 2 | 0.16 | active, pain, use, mouth, occur, hour, education, setup, pt, device |
| | | | 3 | 0.17 | all, comment, up, had, medication, care, pain, more, pt, activity |
| | | | 4 | 0.24 | provided, an, had, more, comment, last, skin, contact, occur, adl |
| | | | 5 | 0.2 | activity, report, use, mg, pain, every, day, hygiene, fall, bowel |
| | 270 | | 1 | 0.16 | active, resident, provider, mg, it, one, daily, problem, status, group |
| | | | 2 | 0.19 | active, tablet, pressure, it, mouth, medication, comment, report, status, one |
| | | | 3 | 0.16 | active, plan, status, comment, pressure, provider, date, support, group, report |
| | | | 4 | 0.22 | active, fall, skin, cm, ml, report, care, mg, call, day |
| | | | 5 | 0.14 | active, resident, mg, one, provider, left, day, pt, skin, ml |
| **High** | 30 | | 1 | 0.12 | tablet, about, pt, date, pain, risk, prn, care, suicide, every |
| | | | 2 | 0.2 | last, call, one, daily, thought, goal, mg, day, refill, skin |
| | | | 3 | 0.42 | up, about, status, day, education, tablet, plan, goal, medication, prn |
| | | | 4 | 0.15 | fall, call, goal, day, treatment, one, risk, education, one tablet, prn |
| | | | 5 | 0.23 | risk, about, report, doe, mg, po, problem, discharge, status, group |
| | 90 | | 1 | 0.13 | active, skin, tablet, disorder, goal, day, prn, medication, blood, risk |
| | | | 2 | 0.21 | assessment, treatment, risk, am, use, care, problem, po, continue, score |
| | | | 3 | 0.23 | am, tablet, self, goal, treatment, date, pt, ml, blood, mood |
| | | | 4 | 0.09 | am, mouth, tablet, denies, pain, last, mood, one, assessment, prn |

| | | | | | |
|---|---|---|---|---|---|
| | | | 5 | 0.17 | active, thought, mood, every, problem, fall, status, level, blood, prn |
| | 270 | BioBERT | 1 | 0.12 | positive, about, mouth, group, intervention, prn, provide, anxiety, urine, up |
| | | | 2 | 0.15 | mood, about, output, one, him, pt, positive, can, urine, up |
| | | | 3 | 0.3 | intake, last, do, po, positive, pressure, care, prn, balance, discussed |
| | | | 4 | 0.17 | nursing, active, output, incontinence, mood, provide, problem, continue, urine, up |
| | | | 5 | 0.29 | may, alcohol, doe, po, hour, skin, pressure, assigned, urine, up |
| | | | 6 | 0.16 | fall, activity, one tablet, level, intake, hour, needed, night, total, use |
| | | | 7 | 0.28 | intake, active, one, group, iv, positive, mouth, can, treatment, urine |
| | | | 8 | 0.17 | doe, active, it, iv, episode, mouth, risk, assigned, total, urine |
| | | | 9 | 0.14 | plan, about, level, may, last, risk, pressure, balance, writer, urine |
| | | | 10 | 0.23 | but, can, pressure, discharge, iv, day, nursing, all, thought, urine |
| | | | 11 | 0.23 | him, activity, ml, last, discussed, pain, prn, but, urine, use |

**Supplementary Table 2: Text+Characteristics Topic Word Importances (cTF-IDF),** with separate topic modeling for reports stratified based on top performing LLM for risk tier and predictive time window, for models which integrated patient characteristics

| Risk Tier | Time Window | Model | Cluster | Suicide Risk Proportion | Top Words |
|---|---|---|---|---|---|
| **Low** | 30 | LongFormer | 1 | 0.18 | active, record, moisture, daily, unit, community, type, every, bowel movement, test |
| | | | 2 | 0.14 | alternate, sleep, service, group, visit, last, th, mouth active, need, twice |

| | | | | | |
|---|---|---|---|---|---|
| | | | 3 | 0.15 | active, treatment, mouth every, imaging, use, medical, set up, mouth active, every day, visit |
| | | | 4 | 0.2 | active, mgdl, mg, choice, office, do, test, eligibility, month, needed |
| | | | 5 | 0.17 | active, screening, month, daily, warfarin, do, unit ref, home, bowel, unit |
| | | | 6 | 0.36 | eligibility, medication, daily, day, screen, every day, collection, mg, condition, total |
| | | | 7 | 0.12 | active, screening, one tablet, day, unit, eval, urine, pt, bowel movement, test |
| | | | 8 | 0.12 | all, th, result unit, goal, up, moisture, specimen, often, how often, warfarin |
| | | | 9 | 0.05 | active, sleep, primary, lab, resident, how, set up, medication, call, sig |
| | | | 10 | 0.15 | range, community care, service, did, choice, dose, self, cm, screening, authorization |
| | 90 | | 1 | 0.11 | active, pressure, right, left, pt, last, provider, twice, may, every |
| | | | 2 | 0.15 | active, pressure, provider, every, mgdl, daily, needed, tablet, medication, last |
| | | | 3 | 0.25 | active, mouth, pt, last, left, daily, pain, up, one tablet, fall |
| | | | 4 | 0.17 | active, right, needed, daily, tablet, education, pain, score, fall, every |
| | | | 5 | 0.18 | daily, pt, call, pressure, every, care, tablet, one tablet, right, left |
| | 270 | BioBERT | 1 | 0.2 | active, one, past, one tablet, every, need, day, mouth, home, last |
| | | | 2 | 0.21 | care, need, mouth, day, risk, it, active, tablet, problem, per |
| | | | 3 | 0.51 | an, group, mouth, one, every, pt, blood, provider, pressure, it |
| | | | 4 | 0.19 | active, need, pain, problem, day, it, daily, mouth, medication, home |
| | | | 5 | 0.18 | active, past, problem, plan, day, needed, every, last, left, home |
| **Med** | 30 | BigBird | 1 | 0.17 | ml, active, pain, consult, scanned document, intervention, iv, last, monitor, but |
| | | | 2 | 0.18 | denies, activity, information, call, provider, medication, lock, education, program, but |
| | | | 3 | 0.21 | following, care, provide, iv, plan, pain, it, pressure, program, every |

| | | | | | |
|---|---|---|---|---|---|
| | | | 4 | 0.17 | pain, about, provider, every, see, click, mouth, medication, disorder, cm |
| | | | 5 | 0.18 | po, military, care, but, doe, problem, mouth, cm, sound, assistance |
| | | | 6 | 0.18 | denies, every, phone, call, provider, pressure, lock, change, treatment, cap |
| | | | 7 | 0.19 | risk, medication, do, active, monitor, following, plan, activity, tablet, assistance |
| | | | 8 | 0.16 | provider, document, pulse, imaging, pressure, required, pt, comment, right, active |
| | | | 9 | 0.19 | medical, about, required, mouth, treatment, click, daily, cm, mood, cap |
| | 90 | | 1 | 0.18 | active, refill, use, medication, mg, last, contact, sig, date, care |
| | | | 2 | 0.18 | active, pain, use, mouth, occur, hour, education, setup, pt, device |
| | | | 3 | 0.18 | all, comment, up, had, medication, care, pain, more, pt, activity |
| | | | 4 | 0.11 | provided, an, had, more, comment, last, skin, contact, occur, adl |
| | | | 5 | 0.17 | activity, report, use, mg, pain, every, day, hygiene, fall, bowel |
| | 270 | LongFormer | 1 | 0.09 | risk, active, left, day, assist, person, pain, alert, pt, up |
| | | | 2 | 0.24 | active, performance, walk, use, provider, iv, every, skin, resident, dressing |
| | | | 3 | 0.22 | activity, occur, walk, up, provided, locomotion, comment, room, safety, day |
| | | | 4 | 0.24 | assist, care, within, up, one tablet, every, activity, plan, physical, pain |
| | | | 5 | 0.18 | active, one tablet, within, use, problem, last, dressing, safety, risk, day |
| | | | 6 | 0.44 | active, mg po, walk, used, problem, occur, device, staff, safety, hand |
| | | | 7 | 0.31 | resident, activity did, device, call, physical, mg, hygiene, daily, dressing, treatment |
| **High** | 30 | | 1 | 0.15 | active, every, treatment, group, use, po, plan, care, tablet, report |
| | | | 2 | 0.12 | active, medication, treatment, group, use, skin, mouth, care, needed, problem |

| | | | | |
|---|---|---|---|---|
| | | 3 | 0.19 | active, every, po, one, report, use, plan, care, risk, tablet |
| 90 | BigBird | 1 | 0.13 | active, skin, tablet, disorder, goal, day, prn, medication, blood, risk |
| | | 2 | 0.09 | assessment, treatment, risk, am, use, care, problem, po, continue, score |
| | | 3 | 0.27 | am, tablet, self, goal, treatment, date, pt, ml, blood, mood |
| | | 4 | 0.11 | am, mouth, tablet, denies, pain, last, mood, one, assessment, prn |
| | | 5 | 0.14 | active, thought, mood, every, problem, fall, status, level, blood, prn |
| 270 | LongFormer | 1 | 0.19 | last, about, stop, every, fall, it, one, thought, supply, sweat |
| | | 2 | 0.16 | nausea, about, sp, mouth, during, level, needle, treatment, tablet, stop |
| | | 3 | 0.28 | it, active, self, mg, intervention, itching, group, thing, symptom, suicide |
| | | 4 | 0.21 | contact, an, status, him, feel, during, minute, stop, skin, report |
| | | 5 | 0.07 | family, about, symptom, expiration, it, fall, po, thought, supply, self |
| | | 6 | 0.25 | him, level, refill, expiration, it, factor, blood, needed, needle, date |
| | | 7 | 0.72 | group, rate, day, intervention, plan, one tablet, contact, blood, but, active |
| | | 8 | 0.28 | did, discharge, status, assistance, group, every, itching, than, sp, rate |
| | | 9 | 0.54 | more, active, mouth, po daily, needed, observation, mg po, thought, symptom, sweat |
| | | 10 | 0.23 | mg po, about, skin, last, intervention, group, needle, thing, stop, status |
| | | 11 | 0.22 | denies, minute, suicide, needle, blood, date, goal, risk, plan, family |

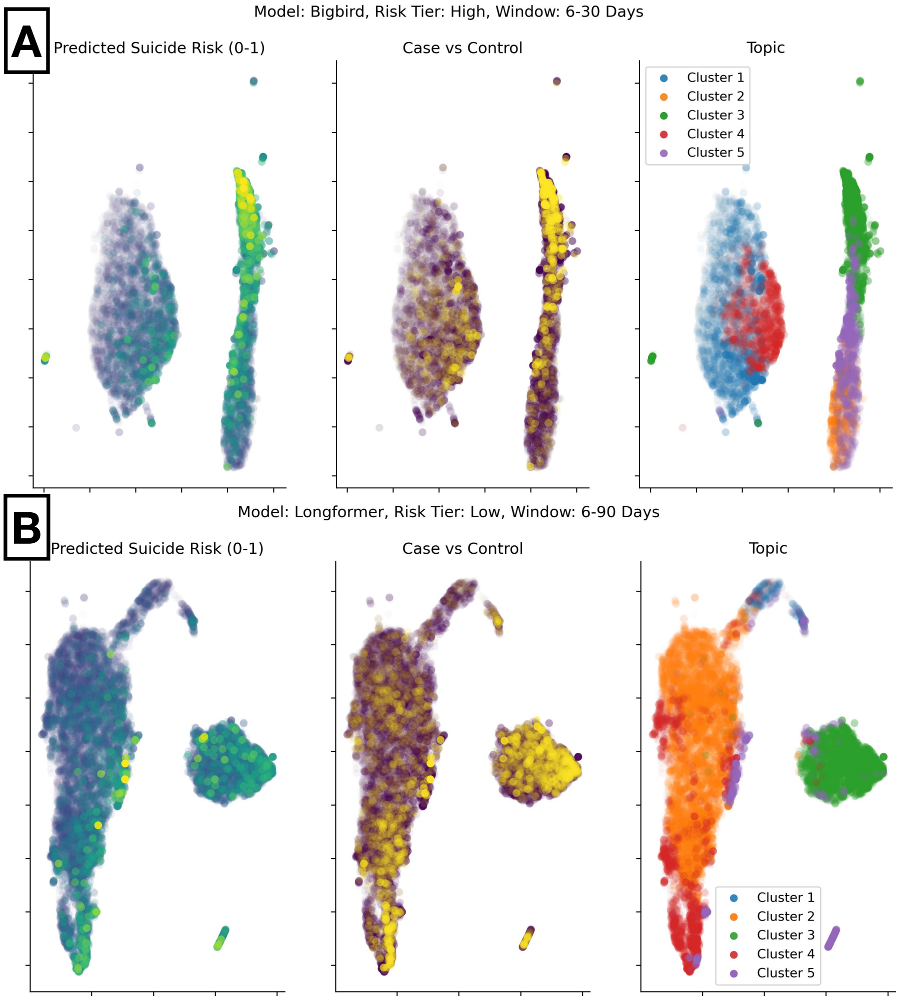


**Supplementary Figure 10: Corpus-level visualization of semantic clusters derived from LLM embeddings.** Two-dimensional UMAP projections of SHAP-derived note embeddings are shown for each model by risk tier and case control status (one combination per row): **A)** the Clinical BigBird model for high-risk patients (6–30 day prediction window) and **B)** the Clinical Longformer model for low-risk patients (6–90 day prediction window). Within each plot each point represents a clinical note, labeled by predicted suicide risk (left), case versus control status (middle), and topic (cluster) assignment (right), respectively. The UMAP dimensions are not interpretable, though closely spaced notes contain similar semantic information, and as such this similarity is used to cluster reports.

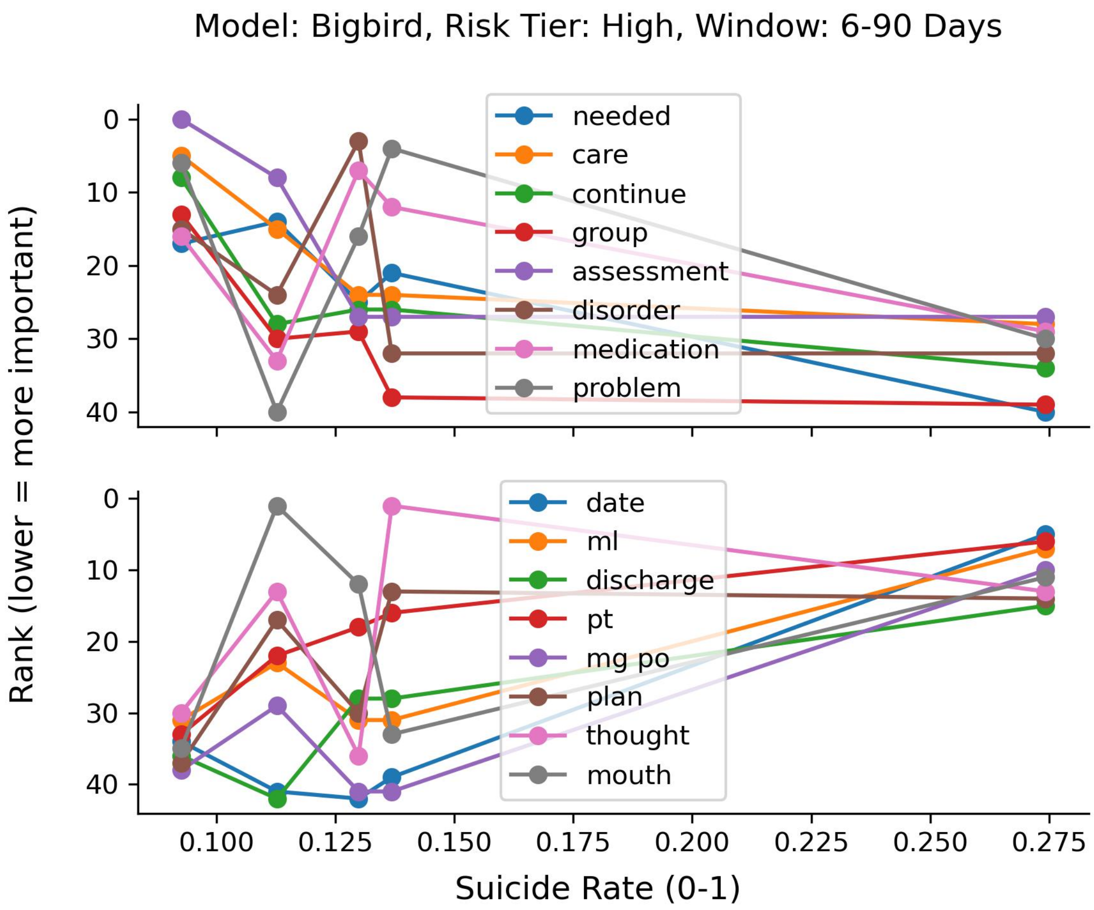


**Supplementary Figure 11: Changes in word relevance across corpus-level topics with increasing suicide risk for the Clinical BigBird model in the high-risk tier (6–90 day prediction window).** The figure shows how the relative importance ranking of selected terms varies across clusters (each x-axis position represents one cluster) with increasing suicide case proportions. The x-axis denotes the cluster-level suicide rate, and the y-axis shows word rank, with lower values indicating higher importance. The top panel highlights terms (e.g., care, assessment, disorder) which decrease in relative importance as suicide risk increases. In contrast, the bottom panel illustrates terms related which become increasingly prominent in higher-risk clusters.

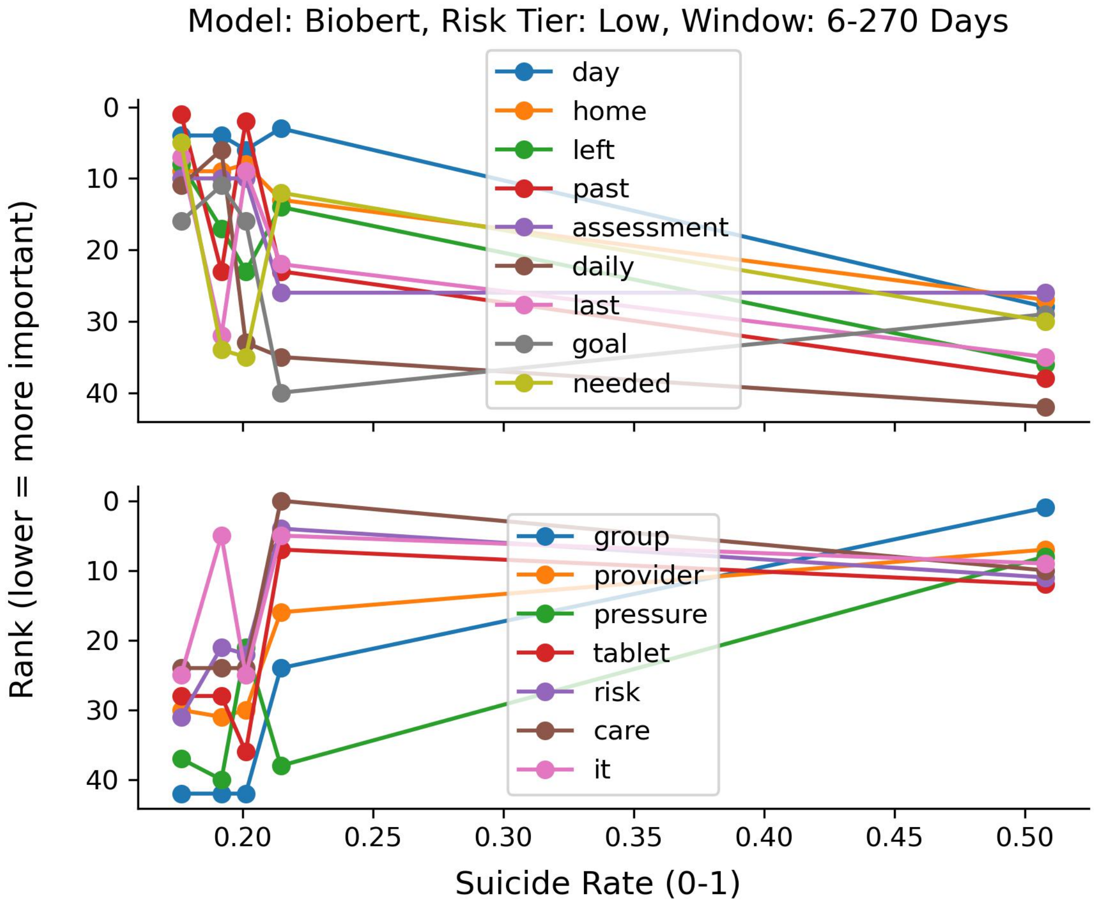


**Supplementary Figure 12: Changes in word relevance across corpus-level topics with increasing suicide risk for the BioClinicalBERT model in the low-risk tier (6–270 day prediction window).** The figure shows how the relative importance ranking of selected terms varies across clusters (each x-axis position represents one cluster) with increasing suicide case proportions. The x-axis denotes the cluster-level suicide rate, and the y-axis shows word rank, with lower values indicating higher importance. The top panel highlights terms (e.g., treatment, pain, medication) which decrease in relative importance as suicide risk increases. In contrast, the bottom panel illustrates terms related which become increasingly prominent in higher-risk clusters.
